# Quantitative proton density mapping with data-driven reception profile correction derived from macromolecular proton fraction

**DOI:** 10.64898/2026.10.13.26365282

**Authors:** Vasily L. Yarnykh

**Author notes:** **Correspondence:** Vasily L. Yarnykh.

## Abstract

**Purpose:** Proton density (PD) and macromolecular proton fraction (MPF) are fundamental MR parameters characterizing water and macromolecular content in tissues. Both parameters can be mapped using a time-efficient MRI protocol, though PD maps require additional corrections for the reception profile (RP) non-uniformity and global scaling. Existing PD mapping methods suffer from uncertainties related to PD normalization to reference tissue. This study aimed to develop a data-driven RP correction method for PD mapping based on MPF as an internal scale.

**Theory and Methods:** One-parameter model linking PD and MPF was derived under assumption of proportionality between total macromolecular volume and proton concentration. This model with a uniform parameter value across brain tissues generates a surrogate PD map, which is used to estimate RP from an uncorrected PD map. The proposed algorithm was tested against a common PD mapping method utilizing bias-field correction and normalization to cerebrospinal fluid. Brain MRI data were acquired at 3T from seven healthy individuals. The protocol included 3D multi-echo *T*_1_-, PD-, and magnetization-transfer-weighted gradient-echo sequences. Parametric maps were reconstructed using the single-point MPF mapping algorithm with data-driven *B*_1_ correction.

**Results:** MPF-based and bias-field corrections yielded similar PD measurements in white and gray matter with no significant bias and remarkably small variability (coefficients of variation <0.6%).

**Conclusions:** MPF-based RP correction enables computationally simple and robust quantitative PD mapping at no additional cost if MPF maps are available. The main advantage of the proposed method is automatic elimination of global scaling that obviates the need for PD normalization.

## Introduction

Proton density (PD) is a fundamental magnetic resonance parameter defining the strength of an MR signal and proportional to water content in tissues [1]. PD demonstrated a promise as a marker of water content for quantitate characterization of brain edema associated with stroke [2–4], tumors [5], inflammation [6–8], and metabolic dysfunction [9]. Besides applications of PD alone, it is used for calculation of certain indexes characterizing macromolecular content, such as macromolecular tissue volume [10] and macromolecular PD [11].

PD maps are typically obtained as a byproduct of relaxation measurements in various *T*_1_ and *T*_2_ mapping methods [1]. However, a number of additional corrections is required to make PD maps usable for water content estimation. These corrections are needed to eliminate residual relaxation effects, transmit *B*_1_ field inhomogeneity, reception profile (RP) non-uniformity, and global intensity scaling [1,12–22]. The variable flip angle (VFA) *T*_1_ mapping method remains one of the most popular approaches for PD quantitation due to fast 3D acquisition and high spatial resolution [15–22]. The VFA method is highly sensitive to *B*_1_ field inhomogeneity and is usually complemented by additional *B*_1_ calibration sequences [15-17,20-22]. Alternatively, *B*_1_ correction can be algorithmically compounded with RP estimation under assumptions of smoothness and linearity of biases caused by transmit and receive fields [18,19]. Within these approaches, either a heavily smoothed low-angle gradient-echo image is used as a proxy for a joint RP and *B*_1_ map [18] or combinations of RP and transmit *B*_1_ are estimated from the linear fit on different voxel subsets [19]. Correction for residual 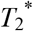 weighting can be either obtained from the fit of multiple gradient echoes [12,15,16,22] or omitted for short TE [17,20,21]. Several techniques were proposed for RP correction including additional image acquired with a whole-body coil used for transmission and reception [13,15], application of bias-field correction algorithms eliminating low-frequency spatial intensity variations [16,17,21,22], reconstruction of RP based on an approximated linear relationship between inverse PD and *R*_1_=1/*T*_1_ [16,19–21], and RP estimation from individual channel images for multi-channel coils [10,20,21]. While these approaches have their own advantages and shortcomings, their common limitation is the need for normalizing a PD map to the standard value in reference tissue (either cerebrospinal fluid (CSF) [16] or white matter (WM) [17]) to correct for global scaling. The rescaling procedure is associated with uncertainties related to tissue segmentation and may be confounded by the presence of lesions or diffuse pathological changes [16,21].

The single-point macromolecular proton fraction (MPF) mapping method [23,24] incorporates two VFA images and one magnetization transfer (MT)-weighted image to reconstruct MPF maps. In combination with the recent data-driven algorithm [25], MPF, *T*_1_, and PD maps produced by the single-point method can be inherently corrected for transmit *B*_1_ inhomogeneity without additional *B*_1_ mapping sequences. While MPF gained substantial popularity over past years as a quantitative myelin biomarker [26], the capabilities of fast MPF mapping protocols expand beyond MPF itself and offer a potential for more comprehensive characterization of tissue microstructure including measurements of water content and associated quantities [27,28]. To uncover this potential, the author presents a new algorithm for accurate and robust PD mapping with automatic RP correction derived from transformed MPF maps. An important feature of the proposed algorithm is simultaneous elimination of both RP non-uniformity and scaling in a single step substantially simplifying the computational procedure.

## Theory

### Surrogate PD estimation from MPF

We follow the two-component tissue model approximating tissue volume (V) as a sum of water (V_w_) and macromolecular (V_m_) volumes [10]. In terms of volume fractions,

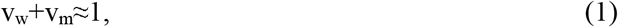

where v_w_=V_w_/(V_w_+V_m_) and v_m_=V_m_/(V_w_+V_m_). PD is defined as a molar concentration of water protons in tissue normalized to the concentration of protons in pure water and expressed in percentage units (pu) [1]:

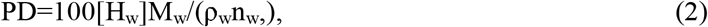

where [H_w_] is the molar concentration of water protons in tissue, M_w_=18 g/mol is the molar mass of water, n_w_=2 is the number of protons in the water molecule, and ρ_w_=993.4 g/l is the water density at 37°C. It is easy to show that v_w_ is equivalent to tissue PD, as defined above. Formally expressing the mass of water (m_w_) via volume and density, m_w_=V_w_ρ_w_, the concentration [H_w_] is given by

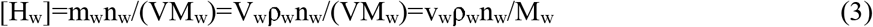

After substituting (3) into (2),

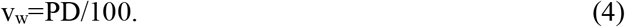

By analogy with Eq. (3), concentration of macromolecular protons can be formally expressed as

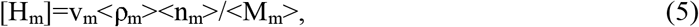

where the densities ρ_m_, numbers of protons per molecule n_m_, and molecular weights M_m_ are averaged over all macromolecular compounds in tissue. Concentrations of water and macromolecular protons are related to the experimentally measured MPF value according to the MPF definition [23,27]:

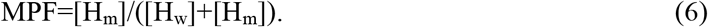

Substituting [H_w_] from Eq. (3) and [H_m_] from Eq. (5), Eq. (6) is transformed to the equality:

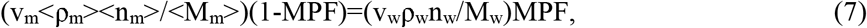

from which, after replacing v_w_ with PD from Eq. (2), v_m_ can be expressed as

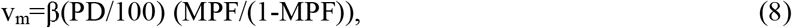

where β is a generally unknown proportionality coefficient absorbing all coefficients in Eq. (7):

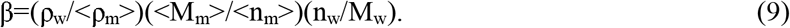

Substituting (4) and (8) into (1) and solving for PD,

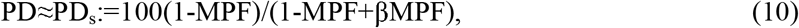

where PD_s_ is defined as a surrogate PD value derived from MPF and approximating actual PD. An appropriate choice of the constant β in Eq. (10) and the feasibility of using a uniform β value across different tissues are investigated below.

### Data-driven estimation of the reception profile

In the single-point MPF mapping method [23,24], a spoiled gradient-echo sequence is used to obtain PD-weighted (PDw), *T*_1_-weighted (T_1_w), and MT-weighted (MTw) images with signal intensities

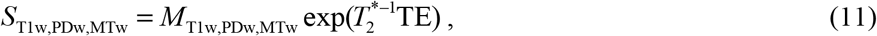

where the magnetization weighting factors *M*_T1w_ and *M*_PDw_ corresponding to flip angles α_T1w_ and α_PDw_ are given by

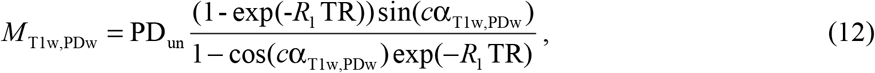

where *c* is the *B*_1_ scaling coefficient, and uncorrected PD (PD_un_) is factored as a product of the global receiver scaling constant *G*, RP, and corrected PD (PD_cor_):

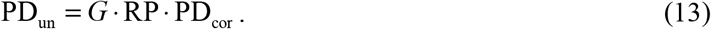

For the MTw image, the weighting factor

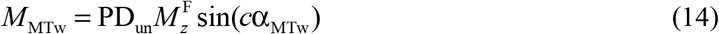

is expressed via the two-pool longitudinal magnetization vector 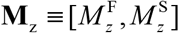 with elements corresponding to the free and semisolid pool and computed by the pulsed steady-state matrix MT equation detailed elsewhere [23]. MPF is obtained by fitting the matrix MT model with the MTw image and *T*_1_ and PD_un_ maps as input data. *B*_1_ non-uniformity can be corrected using either an actual *B*_1_ map [29] or the data-driven algorithm [25] deriving coefficients *c* from uncorrected MPF and *T*_1_.

MPF is inherently insensitive to the reception profile. If Eq. (10) provides sufficiently close approximation of actual PD, it can be used to estimate the product *G*·RP as the ratio of the measured PD_un_ to PD_s_ (*G*·RP≈PD_un_/PD_s_). In combination with an appropriate low-pass filter to suppress residual tissue-dependent variations, the *G*·RP image obtained using this procedure can be applied to an uncorrected PD map to simultaneously correct for both RP non-uniformity and global scaling, thus eliminating the step of PD normalization. In the formal notation, treating images as 3D matrices, the proposed algorithm can be written as

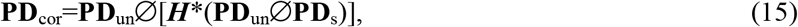

where ***H*** is the filter kernel, the symbol “*” denotes convolution, and the symbol “Ø” denotes Hadamard (element-wise) division.

### Error propagation

Errors in the described algorithm may propagate to the corrected PD via the reconstructed RP from either a bias in the parameter β or uncertainties in MPF. For the purpose of error analysis, it is convenient to present the RP calculation for an arbitrary voxel according to Eqs. (10) and (15) as

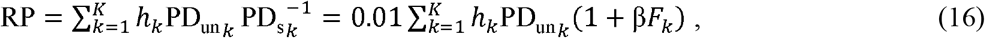

where *h*_*k*_ are the elements of the kernel matrix ***H***, summation is performed over all *K* voxels convolved with the kernel, and MPF is expressed via the pool size ratio *F*: *F*_*k*_=MPF_*k*_/(1-MPF_*k*_). For brevity, the sums are presented in one dimension. Taking into account that ∂PD_cor_/∂RP= −PD_un_ /RP^2^ = −PD_cor_ /RP, absolute errors in PD_cor_ due to the errors in β and *F* are approximated as

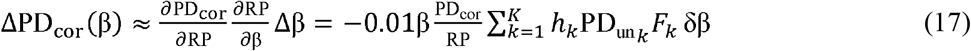

and

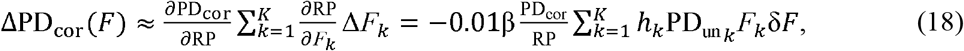

where the symbols Δ and δ denote the absolute and relative errors, and the relative error δ*F* is assumed to be constant for all voxels. Further expressing the sum in Eqs. (17) and (18) via Eq. (16) and assuming that RP is nearly constant across the kernel size (PD_un_ PD_cor_ for any *k*), the equations for relative errors in PD_cor_ are simplified to

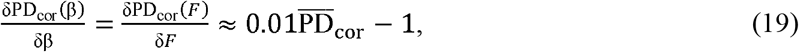

Where 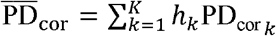 is the weighted average PD_cor_ across the kernel, which reduces to a mean PD_cor_ value if the mean filter (all *h*_*k*_ =1/*K*) is applied. Converting the error in *F* to the error in MPF, Eq. (19) is transformed to

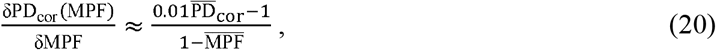

Where 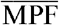 is the mean MPF across the kernel. For a mean filter with a sufficiently large kernel size allowing to encompass substantial proportions of WM, GM, and CSF voxels, 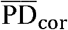 and 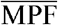 can be approximated by their mean whole-brain values.

## Methods

### Participants

Data were obtained from seven healthy adults (five females; age range 20-60 years) after signing informed consent. The study was approved by the local IRB. Anonymized data from one secondary progressive multiple sclerosis (MS) patient (male, in his 50’s) participated in an earlier published study [30] were reprocessed to assess the performance of the presented method in the presence of brain lesions.

### MRI protocols

Images from six healthy individuals were acquired on a 3T MRI scanner (Ingenia; Philips Medical Systems) with a 32-channel head coil. A customized pulse sequence enabling users’ control of the saturation pulse settings and optimized spoiling [31] was used. The MPF mapping protocol included three spoiled 3D multi-echo gradient-echo sequences: PDw with TR=18 ms and flip angle (FA) 3° (scan time 2 min 50 s); T_1_w with TR=18 ms and FA 20° (scan time 2 min 50 s), and MTw with TR=34 ms and FA 10° (scan time 5 min 20 s). Off-resonance saturation was achieved by the single-lobe sinc-Gauss pulse with offset frequency 4 kHz, effective FA 600°, and duration 12 ms. All images were acquired in the sagittal plane with four-echo unipolar readout (TE_1_/ΔTE=2.3/4.1 ms), FOV=240×240×200 mm^3^, and actual voxel size 1.25×1.25×1.25 mm^3^ interpolated to 0.63×0.63×0.63 mm^3^ after zero-padded Fourier transform. Parallel imaging (SENSE [32]) with acceleration factors 1.8 was used in two phase-encoding directions.

Images from one participant were acquired on the two 3T MRI systems (the above Philips Ingenia and Siemens Magnetom Prisma (Siemens Healthineers)) to demonstrate the applicability of the proposed method to different hardware. The fast single-echo versions of the MPF mapping protocol were implemented on both systems. For the Philips scanner, the 32-channel head coil and the custom sequence referenced above were used. On the Siemens scanner, images were acquired with a 64-channel head coil and an unmodified manufacturer’s 3D spoiled gradient echo sequence. Basic scan parameters were matched between the platforms and were as follows: PDw TR/FA=16 ms/3°; T_1_w TR/FA=16 ms/20°; MTw TR/FA=26 ms/8°; TE=4.6 ms; FOV=240×240×180 mm^3^; acquired voxel size 1.25×1.4×1.4 mm^3^ interpolated to 1.25×1.25×1.25 mm^3^. The single-lobe sinc-Gauss pulse with offset frequency 4 kHz, effective FA 520°, and duration 11 ms was used for MT saturation on the Philips scanner. A standard manufacturer’s Gaussian MT pulse with offset frequency 1.2 kHz, effective FA 500°, and duration 9.98 ms was applied on the Siemens scanner. Manufacturers’ parallel imaging options (SENSE [32] on Philips and GRAPPA [33] on Siemens) were used for acceleration in two dimensions with the factors of 1.5×1.6 on Philips and 2×2 on Siemens, respectively. The total scan time was about 7 min for the Philips and 6 min for the Siemens protocol, respectively.

A subset of images obtained from an MS patient using the previously published protocol [30] was reprocessed in this study. The images were acquired on a 3T Philips (Achieva; Philips Medical Systems) scanner in the axial plane with actual resolution of 1.5×1.5×3.0 mm^3^, TE=2.3 ms, and the following sequence parameters: PDw TR/FA=20 ms/3°; T_1_w TR/FA=20 ms/20°; and MTw TR/FA=43 ms/10°. The MT pulse with the single-lobe sinc-Gauss shape, offset frequency 4 kHz, effective FA 950°, and duration 19 ms was used for off-resonance saturation.

### Image processing and analysis

Unless otherwise stated, reconstruction procedures were carried out using custom C++ software available at https://www.macromolecularmri.org/. Multi-echo images were processed by the single-exponential fit yielding *M*_T1w_, *M*_PDw_, and *M*_MTw_ (Eq. (11)). Subsequent reconstruction steps were carried out on skull-stripped images. Brain masks were created from *M*_PDw_ images using the brain extraction tool [34] implemented in MRIcron software (https://www.nitrc.org/projects/mricron/). Reconstruction of MPF, *T*_1_, and PD_un_ maps with simultaneous correction of transmit *B*_1_ inhomogeneity was performed using the data-driven algorithm [25].

As a reference RP correction method, the bias field removal approach [22] was adopted. Reference PD maps (PD_ref_) were computed from PD_un_ maps using the N4 algorithm [35] implemented in 3D Slicer software (https://www.slicer.org/) [36]. PD_ref_ maps were normalized to the CSF intensity based on the *T*_1_ threshold *T*_1_≥3700 ms [21].

MPF-based RP correction yielding PD_cor_ maps was performed according to Eqs. (10) and (15) with a 3D trimmed mean filter applied to above-zero voxels in RP images. Optimal values of the coefficient β and filter radius *r* were determined for each individual multi-echo dataset by brute-force minimization of the whole-brain voxel-based coefficient of variation (CV_v_) between reference and corrected PD maps:

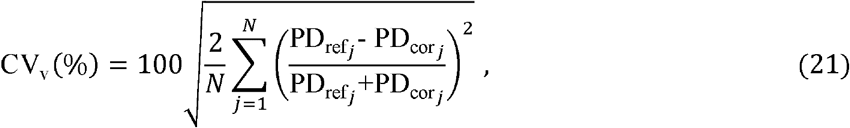

where *N* is the total number of voxels within the brain mask. Then individual β and *r* minimizing Eq. (21) were averaged across all subjects, and all datasets were reprocessed with the uniform mean β and *r* values to compare PD_cor_ and PD_ref_ in brain tissues. Whole-brain WM and gray matter (GM) masks were created by segmentation of MPF maps using FSL software [37,38] (FMRIB Software Library; www.fmrib.ox.ac.uk/fsl/) as detailed elsewhere [24]. Additionally, an estimate of the coefficient β was mapped for illustrative purposes from MPF and PD_ref_ maps:

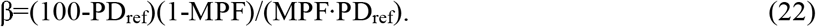

To assess possible errors in PD correction by the MPF-based method, MPF variability was simulated by applying a constant scaling factor to the actual MPF map. PD_cor_ values were measured in WM and GM across a range of MPF scaling factors, β values, and filter radii.

To investigate the applicability of the described method to data obtained without correction of the 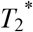 decay, four single-echo datasets from one subject corresponding to each TE were processed separately. The PD measured without compensation of the 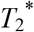 effect is referred below to as PD*. Similarly, PD* maps were reconstructed for the aforementioned single-echo protocols. All reconstructions of PD*_cor_ maps were carried out with a single β value determined as detailed above. Parametric maps obtained on the two MRI platforms from the same subject were aligned to the standard space (Talairach transform) and mutually registered using the rigid-only (6 degrees of freedom) 3D automatic registration procedure implemented in MIPAV software (Medical Image Processing, Analysis and Visualization; https://mipav.cit.nih.gov/) [39].

### Statistical analysis

Agreement between PD values in segmented WM and GM obtained with the MPF-based and reference correction methods was assessed using the Bland-Altman plots and within-subject coefficient of variation (CV). Corrected by the MPF-based method and surrogate PD values were compared with the reference PD values using the Wilcoxon signed-rank test. The statistical significance level (two-sided) was set at *p*<0.05. Data are presented as the mean ± SD.

## Results

### Algorithm optimization

An overview of parametric images produced by the data-driven PD correction algorithm is presented in Fig. 1. The MPF map (Fig. 1A) is used to calculate a surrogate PD map (Fig. 1B). The surrogate PD map shows substantially similar to actual PD (Fig. 1C) tissue contrast. Quantitatively, surrogate PD values were significantly smaller than reference PD in WM (mean PD_s_ = 69.97 ± 1.00 pu, mean PD_ref_ = 72.00 ± 0.90 pu; *P*=0.03) but did not differ significantly in GM (mean PD_s_ = 83.35 ± 0.79 pu, mean PD_ref_ = 83.08 ± 0.73 pu; *P*=0.31). This observation indicates that despite visual similarity, surrogate PD maps cannot be used as the replacement of actual PD maps. The unfiltered RP map (Fig. 1D) computed from uncorrected and surrogate PD maps shows a clear pattern of low-frequency RP non-uniformity superimposed on minor residual tissue-dependent variations. Tissue-related variability is effectively removed after filtering (Fig. 1E). The map of the coefficient β (Fig. 1F) calculated by Eq. (22) shows rather uniform brain tissue appearance without visible distinctions between WM and GM. The only variations observed in the β map are elevated values associated with CSF-tissue interfaces and a slight reduction in the globus pallidus (Fig. 1F). The former can be explained by the uncertainty of β calculation for low MPF values, while the latter is likely due to the residual 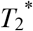 effect in iron-rich regions propagating into the reference PD map.

**FIGURE 1.**
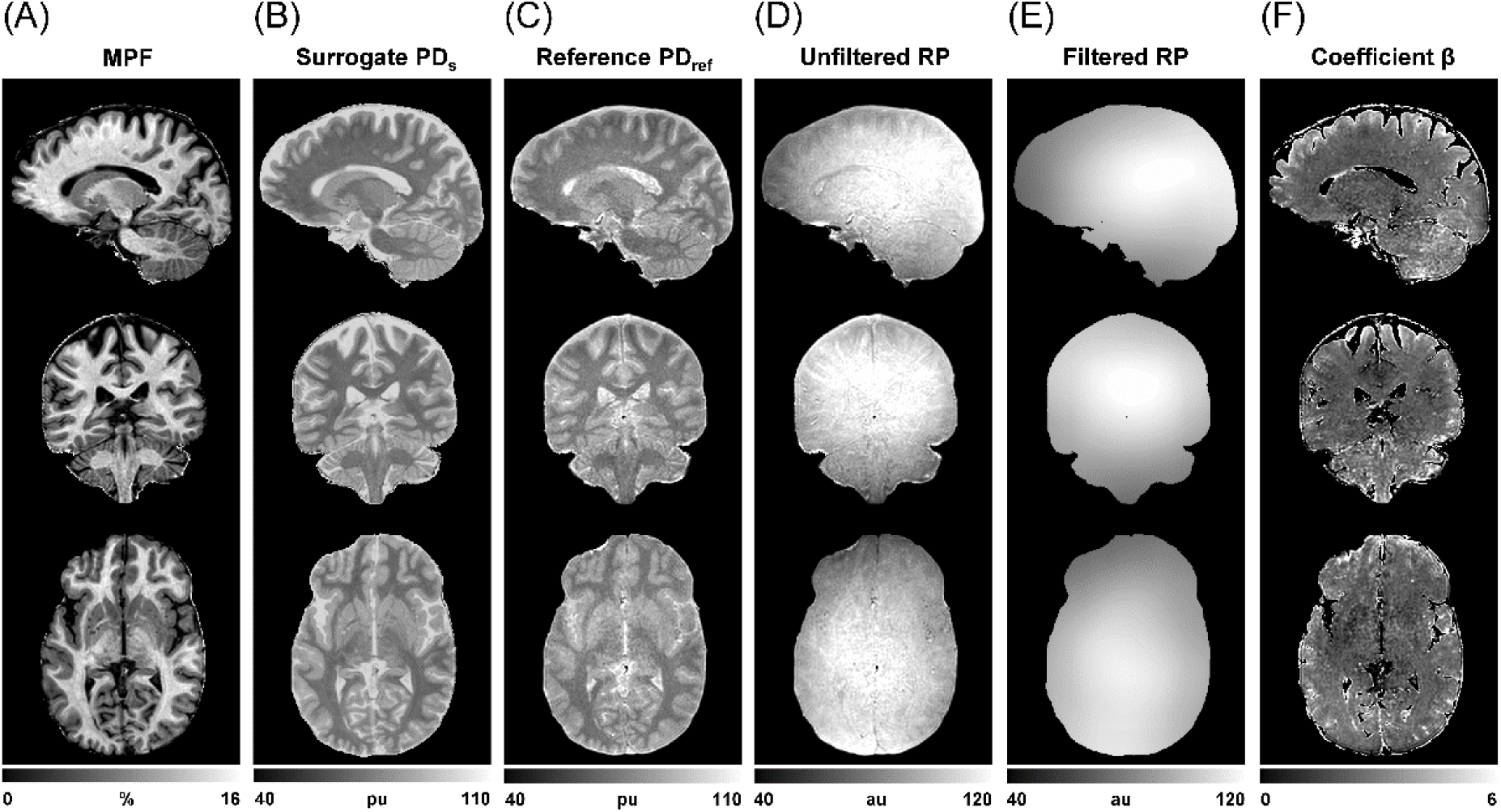
Principle of the data-driven MPF-based RP correction method. (A) MPF map. (B) Surrogate PD map calculated from the MPF map with β = 2.8. (C) Reference PD map corrected using the bias-field removal method. (D) Map of the RP computed from the uncorrected and surrogate PD maps before applying a filter. (E) The same RP map smoothed using the 3D trimmed mean filter with *r* = 22 mm. (F) Map of the coefficient β calculated from the MPF and reference PD maps with a threshold of MPF>1% to avoid division by near-zero values.

The optimal β value was found by minimizing the voxel-based CV between the reference and MPF-corrected PD maps (Fig. 2A). For all individual datasets, the optimal β values (minimum positions) appeared in a close range between 2.6 and 3.1 with the mean β=2.8±0.2 and mean CV_v_=2.3±0.5%. A similar approach was used to find an optimal filter radius (Fig. 2B). Filtering dramatically reduced variation between the reference and MPF-corrected PD maps. The dependence of CV_v_ on the filter radius demonstrated a flat minimum around *r* = 22±1 mm (range 20-24 mm). Based on these findings, the values of β=2.8 and *r*=22 were subsequently used unless otherwise stated.

**FIGURE 2.**
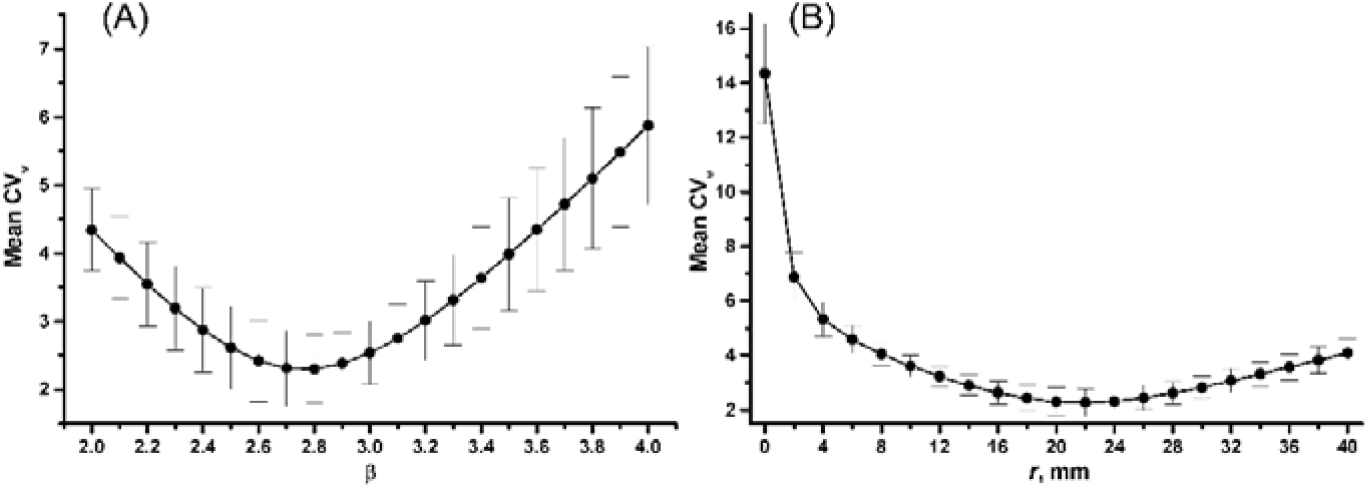
Mean voxel-based coefficient of variation (CVv) between PD maps corrected using the reference and MPF-based methods plotted as a function of the coefficient β (A) and radius *r* of the 3D trimmed mean filter (B). CV_v_ values are averaged across 6 individual datasets. Error bars represent SD. In plot (A), *r* = 22 mm. In plot (B), β = 2.8.

### Agreement with the reference method

Figure 3 compares the reference and MPF-based correction methods in an example dataset. An uncorrected PD map (Fig. 3A) shows obvious coil-related intensity variations. The PD maps corrected using the bias-field and MPF-based methods (Fig. 3B,C) appear very similar. The effect of RP correction is further highlighted by the difference between corrected and uncorrected PD maps (Fig. 3D), which reflects both RP non-uniformity and tissue-dependent variability caused by the multiplicative bias. The difference between the two RP-corrected maps (Fig. 3E) shows only a low-frequency pattern with minor variations. Agreement between the two correction techniques is further evidenced by similarity of the RP maps computed using the bias-field (Fig. 3F) and MPF-based (Fig. 3G) algorithms.

**FIGURE 3.**
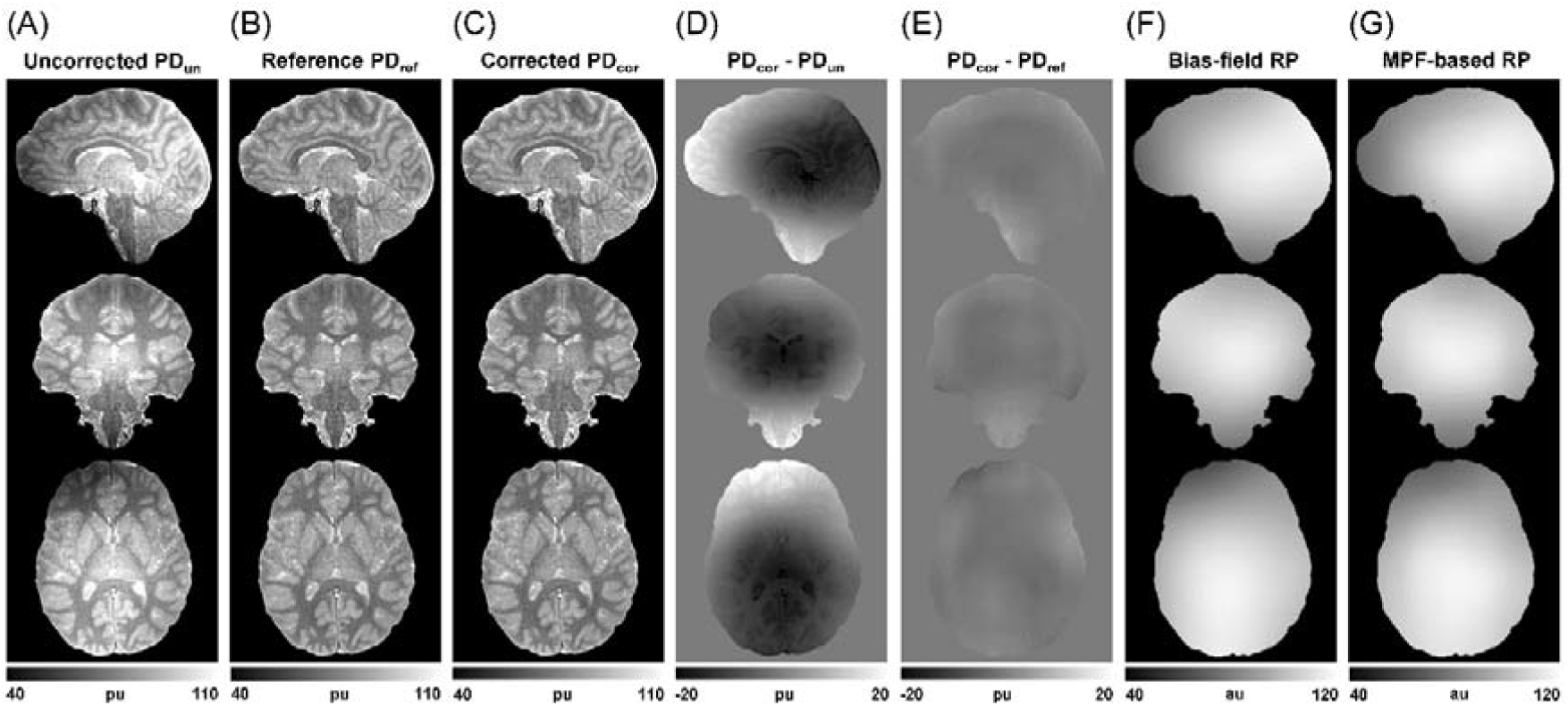
Example dataset illustrating agreement between MPF-based and reference RP correction methods. (A) Uncorrected PD map. (B) Reference PD map corrected using the bias-field removal method. (C) PD map corrected using the MPF-based method. (D) Difference between MPF-corrected and uncorrected PD maps. (E) Difference between MPF-corrected and reference PD maps. (F) RP map produced by the reference bias-field correction algorithm. (G) RP map computed using the MPF-based algorithm.

The Bland-Altman plot characterizing agreement between corrected PD measurements in segmented brain tissues is presented in Fig. 4. Corresponding statistical parameters are listed in Table 1. Overall, MPF-based and bias-field corrections yielded very close PD measurements in WM and GM with no significant bias, narrow limits of agreement (<±1.6 pu), and extremely small variability (CV<0.6%).

**TABLE 1.** Comparison between PD values corrected using the reference and MPF-based methods.

| Value | WM | GM |
| --- | --- | --- |
| Reference method, $PD_{ref}$ (pu) | $72.00 \pm 0.90$ | $83.08 \pm 0.73$ |
| MPF-based method, $PD_{cor}$ (pu) | $71.66 \pm 0.45$ | $83.43 \pm 1.05$ |
| Mean PD (pu) | $71.83 \pm 0.66$ | $83.26 \pm 0.85$ |
| Bias (pu) | $0.32 \pm 0.49$ | $-0.36 \pm 0.61$ |
| Significance of bias ($P$) | 0.22 | 0.31 |
| Limits of agreement (pu) | -0.65; 1.29 | -1.55; 0.83 |

**FIGURE 4.**
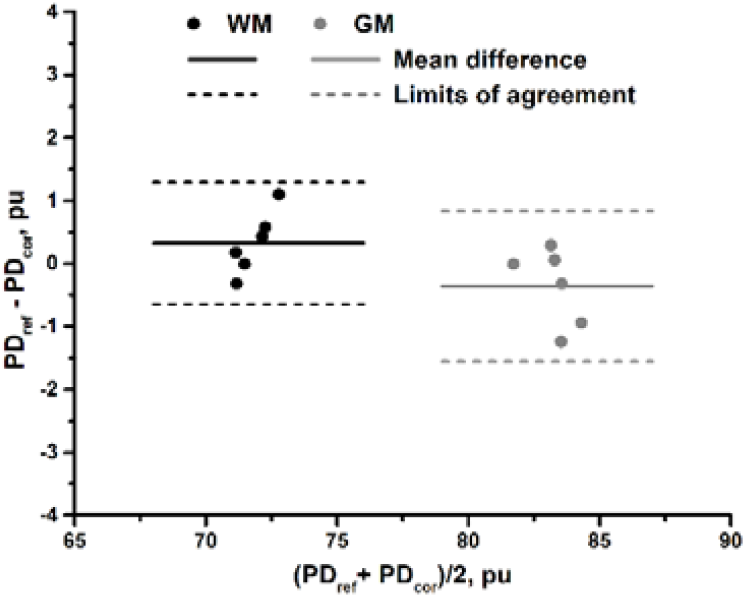
Bland-Altman plot comparing PD values in WM (black dots) and GM (gray dots) measured from the PD maps corrected using the reference and MPF-based methods. Solid and dashed lines correspond to the mean difference and limits of agreement, respectively.

### Error analysis

Relative errors in the corrected PD caused by variation of the input MPF scaling factor, constant β, and filter radius based on a single dataset are plotted in Fig. 5. The relative errors in PD_cor_ as functions of relative errors in MPF (Fig. 5A) and β (Fig. 5B) appear nearly linear and almost identical for WM and GM. The errors computed from the real data demonstrate excellent agreement with the theoretical predictions calculated using Eqs. (19) and (20) for the approximate whole-brain values of 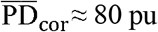 and 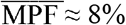. Notably, relative errors in PD_cor_ are characterized by an about five-fold-narrower range than the input errors in either MPF (Fig. 5A) or β (Fig. 5B). The radius of the mean filter also produces a relatively minor effect on the PD_cor_ estimates (Fig. 5C). Specifically, variations of PD_cor_ do not exceed ±1% for a wide range of filter sizes (*r*≈15-35 mm). For smaller filter kernels, errors increase due to propagation of incompletely suppressed tissue-dependent variations in RP, while larger filter radii may cause oversmoothing of RP and, therefore, incomplete non-uniformity correction in the resulting PD_cor_ map.

**FIGURE 5.**
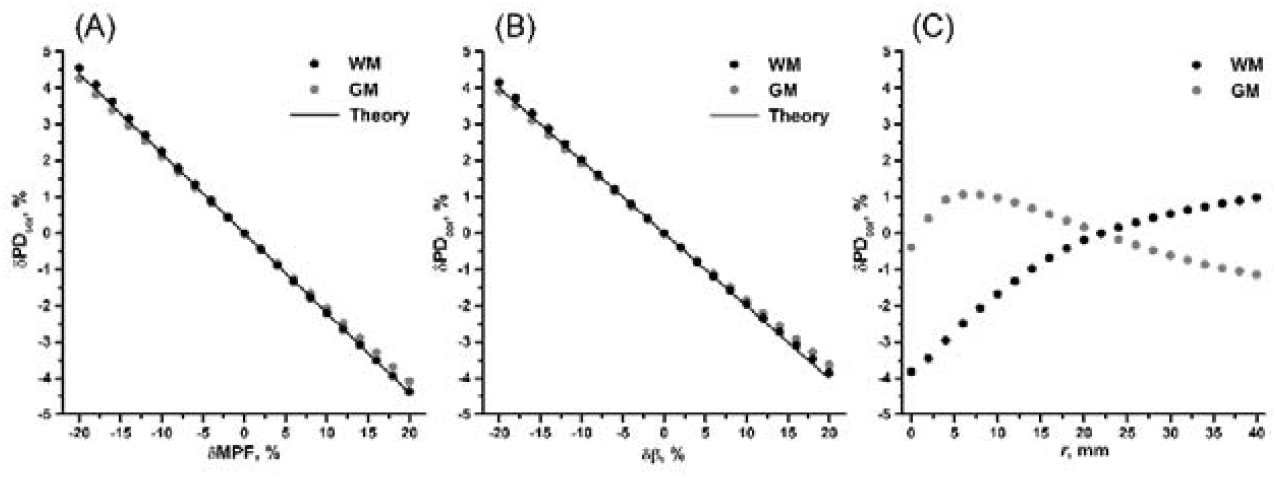
Relative errors in PD corrected using the MPF-based method plotted as functions of a relative error in MPF (A), a relative error in the coefficient β (B), and a radius of the trimmed mean filter *r* (C). Dots correspond to the measurements in segmented WM (black) and GM (gray) taken from the PD_cor_ maps reconstructed with a variable MPF scaling factor (A), β (B), or *r* (C). Errors are calculated relative to the PD values obtained with the unmodified MPF map, β = 2.8, and *r* = 22 mm. The straight lines in plots (A) and (B) correspond to the theoretical model (Eqs. (19), (20)) with 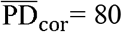 and 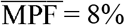

### Compensation of 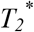 decay

Example reconstructions of PD* maps at variable TE using the MPF-based and reference methods are presented in Fig. 6A. The difference between these reconstructions progressively increases with an increase in TE due to a decrease in the reference PD* values. This effect is highlighted by the whole-brain PD* histograms for the reference method (Fig. 6B), which exhibit a marked shift to lower values with TE elongation. The subtraction images between the 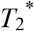-corrected PD_cor_ map and TE-dependent PD*_cor_ maps (Fig. 6A) demonstrate a different type of behavior. They do not show global intensity changes but reflect amplification of the contrast between WM and GM and enhanced hyperintensity of the iron-rich globus pallidus with an increase in TE. The position of the PD* histograms for the MPF-based correction method remains relatively stable even at the longest TE (Fig. 6C), while the TE effect is manifested as slightly increased separation between the WM and GM peaks and noise-related histogram broadening. These observations indicate that MPF-based correction partially compensates for the effect of 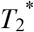 decay and markedly reduces corresponding systematic errors in PD* as compared to the reference method.

**FIGURE 6.**
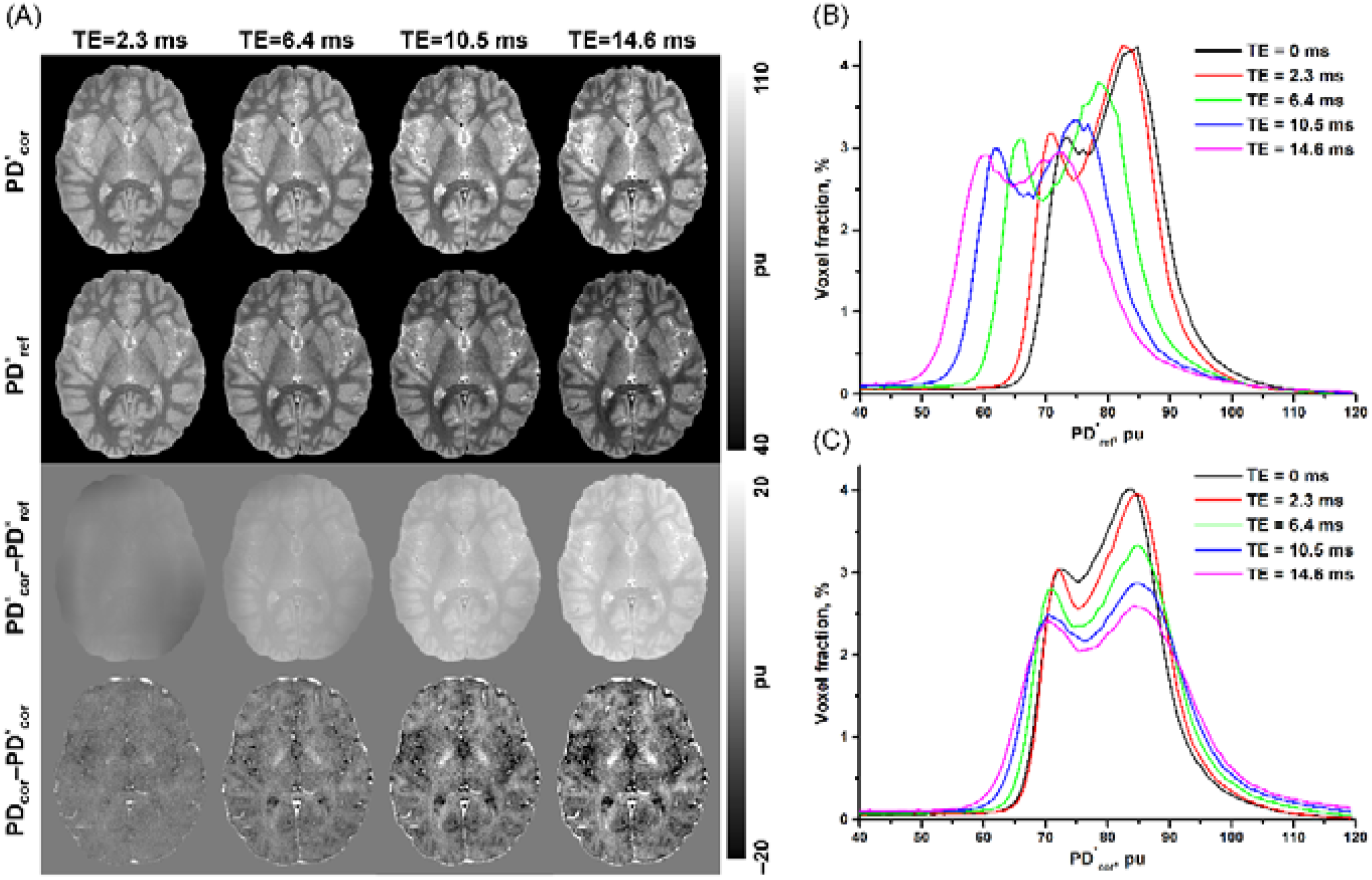
Dependence of PD* corrected using the MPF-based and bias-field methods on TE. (A) Representative cross-sections of PD*maps reconstructed for TE = 2.3, 6.4, 10.5, and 14.6 ms with MPF-based correction (top row) and reference bias-field correction (second row); their difference images (third row); and subtraction images between the PD map corrected for 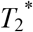 decay by the multi-echo fit and the TE-dependent PD* maps for the MPF-based method (bottom row). (B) Whole-brain PD* histograms after bias-field removal for each of the above TE. (C) Whole-brain PD* histograms after MPF-based correction for each of the above TE. Histograms corresponding to TE = 0 ms in panels (B) and (C) refer to 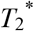-corrected PD.

### Comparison between the two MRI systems

Evaluation of the MPF-based PD correction on the 3T Siemens and Philips scanners based on the data from the same participant is presented in Figs. 7 and 8. Uncorrected PD maps (Fig. 7A,B) and RP (Fig. 7C,D) demonstrated divergent inhomogeneity patterns related to different receiver coil designs. Notably, the Siemens system is characterized by a highly non-uniform reception profile with an about five-fold inhomogeneity range (Fig. 7C). To avoid RP oversmoothing, the filter radius for the Siemens dataset was empirically adjusted to 5 mm. MPF maps showed consistent appearance on both imaging platforms (Fig. 7E,F). Correction using the MPF-based method resulted in nearly identical PD*_cor_ maps (Fig. 7G,H) with the difference image demonstrating a uniform noise pattern except for the minor edge effects (Fig. 7I). Correction using the reference bias-field method produced less consistent results (Fig. 7J,K) as reflected by apparent residual inhomogeneity in the difference image (Fig. 7L). Whole-brain PD* histograms (Fig. 8) further highlight the distinctions in the performance of the correction methods. The histograms of PD* corrected using the MPF-based method appear closely aligned, while the histograms of the bias-field-corrected PD* show substantial disagreement. Bias-field correction of the Philips data demonstrates the shift to lower PD* values consistent with the TE effect described above. For the Siemens data, the histogram is shifted to higher PD* values due to incomplete non-uniformity correction, which does not fully restore the signal drop in the center of RP and causes normalization to the spuriously reduced ventricular CSF signal.

**FIGURE 7.**
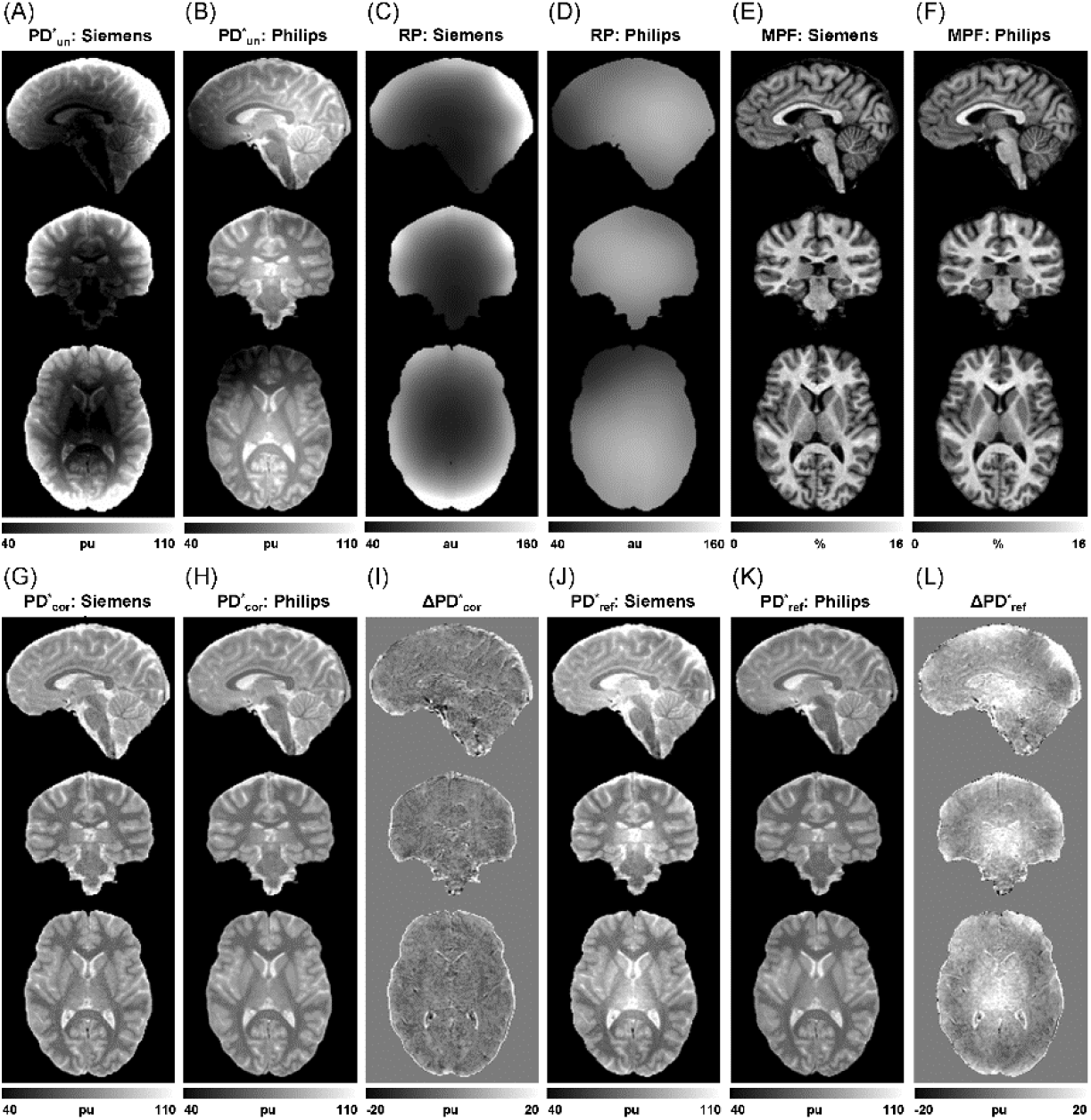
Comparison between the MPF-based and bias-field PD* correction methods applied to the data acquired on the two 3T MRI systems from the same participant: uncorrected PD* maps obtained on the Siemens (A) and Philips (B) scanners; RP reconstructed by the MPF-based method from the Siemens (C) and Philips (D) data; MPF maps obtained on the Siemens (E) and Philips (F) scanners; MPF-corrected PD* maps obtained on the Siemens (G) and Philips (H) scanners and their difference (I); bias-field-corrected PD* maps obtained on the Siemens (J) and Philips (K) scanners and their difference (L).

**FIGURE 8.**
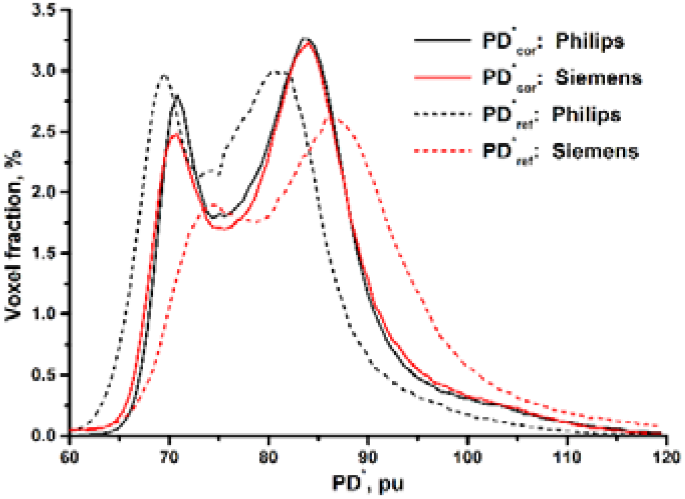
Whole-brain PD* histograms corresponding to the RP-corrected PD* maps obtained from the same participant on the two 3T MRI systems and presented in Fig. 7. Solid and dashed lines represent the MPF-based and bias-field non-uniformity correction methods, respectively. Black and red colors correspond to the datasets acquired on the Philips and Siemens scanners, respectively.

### MS example

Application of the described method to a dataset obtained from an MS patient is illustrated in Fig. 9. The *T*_2_-weighted FLAIR image (Fig. 9A) reveals multiple lesions, which are characterized by a variable extent of demyelination according to the MPF map (Fig. 9B). Most lesions are also clearly seen in the corrected PD map (Fig. 9C). The unfiltered RP map (Fig. 9D) does not demonstrate visible propagation of the contrast features related to lesions. Subtle residual high-frequency variations associated with CSF-tissue interfaces and minor ghosting artifacts are completely eliminated by filtering (Fig. 9E).

**FIGURE 9.**
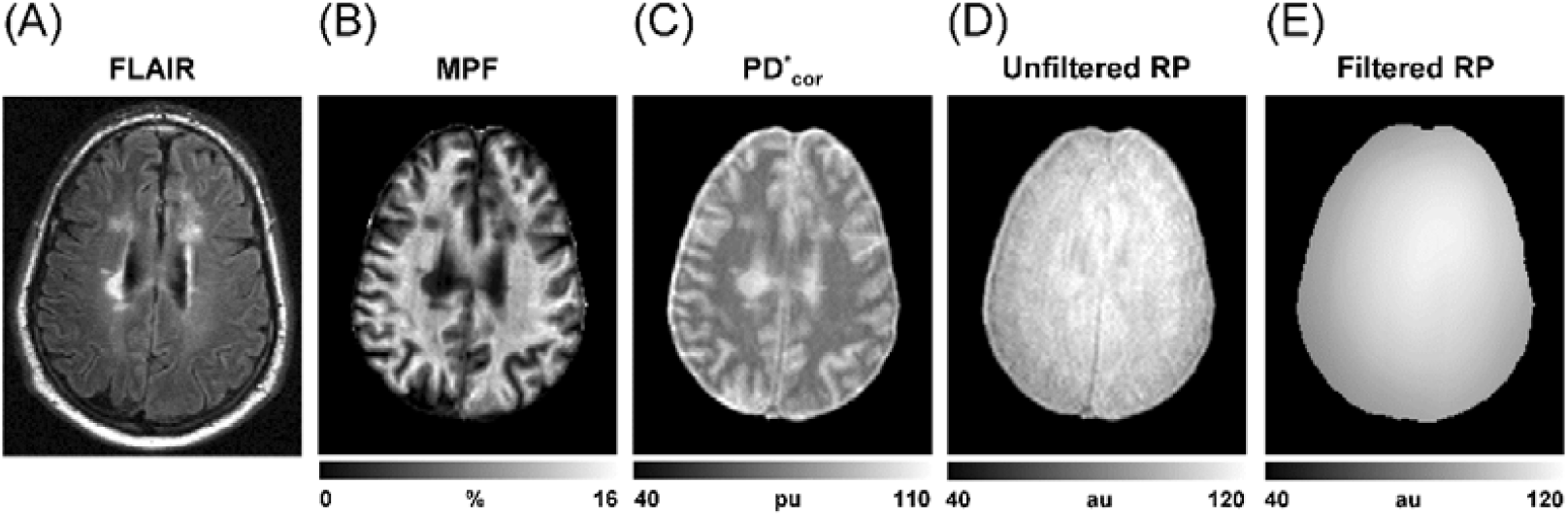
Application of the MPF-based RP non-uniformity correction for quantitative PD* mapping in MS. (A) *T*_2_-weighted FLAIR image used to identify lesions. (B) MPF map depicting the extent of demyelination. (C) PD* map corrected using the MPF-based method. (D) Unfiltered RP map. (E) Smoothed RP map after applying the 3D trimmed mean filter.

## Discussion

This study introduces a novel application of MPF maps as a means for correction of RP non-uniformity and scaling in PD maps. The new PD mapping approach demonstrated excellent agreement with a commonly used bias-field correction technique and produced consistent with the literature PD measurements in normal WM and GM (reviewed in [1]). In combination with the earlier data-driven *B*_1_ correction algorithm [25], the presented technique makes single-point MPF mapping [23,24] a comprehensive time-efficient technology for quantitation of brain water and macromolecular composition.

A key advantage of the described method is elimination of the intensity rescaling step from the PD map reconstruction pipeline. Besides uncertainties related to tissue segmentation, rescaling is a major source of PD quantitation errors, since any error in the global normalization constant translates into the same error in the measured PD on the percentage basis. The MPF-based method offers a more favorable error profile with relative errors in PD being about five-fold smaller than relative errors in either MPF or coefficient β. Another important practical consequence of the elimination of rescaling is a partial compensation of the 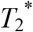 decay. PD mapping techniques often use single-echo acquisition to shorten scan time and simplify processing [17,20,21]. PD* maps obtained from single-echo data are typically subjected to the bias caused by 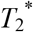 relaxation. The methods based on PD normalization handle the 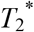 effect differently from the proposed method, especially if normalization is based on the CSF signal. Since 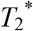 of CSF is considerably longer compared to tissues, 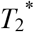 decay progressively propagates into PD* maps with TE elongation. In contrast, the MPF-based method implicitly treats the 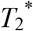 decay as a part of global RP scaling, thus eliminating the common for all tissues portion of this effect. As a result, MPF-based correction produces PD* values being substantially closer to the actual PD as compared to the rescaling-based methods.

The proposed method relies on a simple single-parameter model describing a relationship between the two fundamental quantitative MRI parameters: PD and MPF. The measured value β=2.8 aligns well with the reference literature values of PD [1] and MPF [23,40] in normal human brain tissues. Particularly, for WM with MPF of 12-14% [23,40] and PD≈70 pu [1], β can be estimated according to Eq. (22) in a range of 2.6-3.1. Similarly, for GM with MPF of 6-7% [23,40] and PD≈84 pu [1], β falls in a range of 2.5-3.0. A relatively uniform distribution of the parameter β in the brain parenchyma justifies the use of a constant whole-brain β value in the data-driven RP estimation algorithm. However, this model should be viewed as an approximation disregarding certain features of tissue composition, such as fat and mineral content. The presence of demyelinating brain lesions does not affect possible applications of the proposed technique. As seen from the presented MS example, the effect of propagation of the lesion contrast into the RP map appeared practically negligible. While extensive discussion of prospective clinical applications of the described method is beyond the scope of this study, potentially problematic areas may include large lesions with tissue properties substantially departing from Eq. (10). These may include calcified tissue, hemorrhage, or some brain tumors. The use of MPF-based PD correction in the presence of such lesions may need to be combined with lesion masking and RP interpolation [16].

Filtering is an important component of the proposed algorithm intended to suppress residual tissue-dependent features but preserve low-frequency variations in RP. While a variety of filters can be used for this purpose, a simple trimmed mean filter showed good performance in this study. An optimal filter radius depends on the coil design. This parameter should be chosen to provide sufficient smoothness of RP but avoid over-smoothing, which would cause incomplete non-uniformity correction in PD maps. Based on visual inspection of unfiltered RP maps obtained in this study, intensity variations associated with residual tissue contrast typically appear within the ±5% range. This observation suggests an empirical rule that the filter radius should match an about 10% intensity change due to the RP intensity gradient. For example, the Siemens 64-chanel coil (images are presented in Fig. 7) provides an about 20% per centimeter average intensity gradient, which translates into the 5 mm filter radius. Similarly, for the Philips 32-chanel coil, an average gradient of about 5% per centimeter leads to the filter radius of about 20 mm, which perfectly agrees with the value found via optimization (Fig. 2). It is also important to note that adjustment of the filter radius does not require high accuracy, since possible errors associated with the filter size are rather minor, as exemplified in Fig. 5. In a practical situation, filter adjustment can be performed once for a specific coil based on visual inspection of RP obtained in trial computations.

Although bias-field correction has been chosen in this study as the reference method due to its simplicity and broad availability, this approach is prone to certain limitations. Suboptimal performance of bias field removal was observed in the presence of very strong RP non-uniformity. The earlier literature [20] demonstrated that bias-field correction is less accurate for PD quantitation than *R*_1_-based correction. As noted above, the reference technique used in this study is also more sensitive to errors caused by 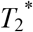 decay. While the bias-field and MPF-based methods preformed equally well on the multi-echo datasets acquired using a coil with a relatively smooth RP and preprocessed with the 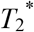 fit, their distinctions became apparent in the experiment involving the two MRI systems. This experiment demonstrated a substantially improved performance of MPF-based correction enabling close agreement between the PD* maps obtained with different MRI hardware and protocols.

Conceptually, the proposed method has some similarity to the earlier approach [16] based on calculation of “pseudo-PD” values from *T*_1_. However, the use of MPF in this context offers several advantages. Particularly, the model linking PD and MPF has only one parameter, whereas association between PD and *T*_1_ is described by two parameters [41]. The dependence of PD on MPF given by Eq. (10) is more biophysically plausible, since both quantities in brain tissues are primarily determined by macromolecular content. Conversely, the corresponding dependence for *T*_1_ can be confounded by paramagnetic ions (mainly iron) [42]. Due to minor tissue-dependent variability of the coefficient β, RP can be computed over the whole brain in the presented method. The *T*_1_-based approach usually utilizes only a subset of voxels followed by interpolation of the entire RP [16,19–21]. This makes the correction procedure more complex and renders the need for subsequent PD rescaling. Since *T*_1_ depends on magnetic field, coefficients of regression between 1/PD and *R*_1_ need to be experimentally determined for each field strength. In contrast, both PD [43] and MPF [44,45] are independent of magnetic field, thus making Eq. (10) with the standard β value universally applicable to any field strength.

The numerical value of the coefficient β found in this study provides useful insights into a relationship between the total macromolecular content and amount of macromolecular protons causing the MT effect. Based on β=2.8 and assuming <ρ_m_>≈1160 g/l (calculated for 75% water content as detailed in Supporting Information Table S1), the ratio <M_m_>/<n_m_> can be estimated from Eq. (9) as 29 g/mol. Direct estimate of <M_m_>/<n_m_> can be obtained from brain tissue elemental composition data [46]. After recalculating for dry weight (Supporting Information Table S1), actual <M_m_>/<n_m_>≈10 g/mol. The observed mismatch suggests that the semisolid proton pool probed by MPF constitutes about 1/3 of all macromolecular protons in brain tissues, whereas the remaining 2/3 appear excluded from magnetization exchange. The discrepancy between the stoichiometric and observed macromolecular proton pool size may reflect the fact that tissue represents a complex mixture of many types of macromolecules with substantially different cross-relaxation properties. The studies of individual lipid components of myelin and other plasma membranes [47–50] indicate that longitudinal relaxation and the MT effect in brain tissues are dominated by certain classes of lipids. The excluded protons likely originate from mobile macromolecules (such as globular proteins) and side chains, which occupy a portion of the tissue volume but do not form ordered supramolecular structures. Furthermore, even within a single class of membrane lipids, a complex equilibrium of liquid crystal phases with distinct molecular order may exist. Their proportion may depend on various biochemical factors including concentration, temperature, pH, and the presence of other constituents [47–50]. Conversely, a minor variability of β in brain tissues found in this study suggests that the tissue dependences of MPF and PD are mainly driven by the bulk sizes of the macromolecular and water proton pools rather than the properties of individual macromolecular components.

## Conclusions

This study introduces a new method for RP non-uniformity correction in quantitative PD mapping based on the approximate theoretical relationship between PD and MPF. The key advantage of the proposed RP correction algorithm is automatic elimination of global intensity scaling that obviates the need for PD normalization to any reference tissue. MPF-based RP correction enables a computationally simple and robust PD mapping technique at no additional cost if MPF maps are available.

## Data Availability

All data produced in the present study are available upon reasonable request to the author

## Funding

This study was supported by the National Institutes of Health grant R01NS136227. Software for MPF map reconstruction was developed under support of the National Institutes of Health grant R24NS104098.

## Data Availability Statement

The data that support the findings of this study are available from the author upon reasonable request. Software code containing implementation of the described algorithm is available at https://www.macromolecularmri.org/downloads/software (Software for MPF, T1, R1, and PD map reconstruction, v. 3.0).

## Notes

### Competing Interest Statement

The authors have declared no competing interest.

### Author Declarations

IRB of the University of Washington gave ethical approval for this work.

