## Supporting Information Table S1 for "Quantitative proton density mapping with data-driven reception profile correction derived from macromolecular proton fraction"

**TABLE S1. Brain tissue elemental composition and density according to the literature [46].**

| Property | Wet weight [46] | Dry weight (recalculated)^a^ | Water |
| --- | --- | --- | --- |
| Elements: mass % |  |  |  |
| H | 10.7 | 9.6 | 11.1 |
| O | 71.2 | 18.0 | 88.9 |
| C | 14.5 | 58.0 | - |
| N | 2.2 | 8.8 | - |
| P | 0.4 | 1.6 | - |
| S | 0.2 | 0.8 | - |
| Other elements | 0.8 | 3.2 | - |
| Macromolecules^b^: mass % | 24.2 | 96.8 | - |
| Water: mass % | 75.0 | - | 100 |
| Molar mass per hydrogen atom: g/mol | - | 10.1^c^ | 9 |
| Density: g/l | 1040 | 1160 | 1000 |

^a^Assuming water content of 75% by weight.

^b^Assuming that macromolecules include all C, N, P, and S atoms and non-water O and H atoms.

^c^Excludes other elements, estimate of <M_m_>/<n_m_>.
